# Utrametric diffusion model for spread of covid-19 in socially clustered population: Can herd immunity be approached in Sweden?

**DOI:** 10.1101/2020.07.15.20154419

**Authors:** Andrei Khrennikov

## Abstract

We present a new mathematical model of disease spread reflecting specialties of covid-19 epidemic by elevating the role social clustering of population. The model can be used to explain slower approaching herd immunity in Sweden, than it was predicted by a variety of other mathematical models; see graphs Fig. 2. The hierarchic structure of social clusters is mathematically modeled with ultrametric spaces having treelike geometry. To simplify mathematics, we consider homogeneous trees with *p*-branches leaving each vertex. Such trees are endowed with algebraic structure, the *p*-adic number fields. We apply theory of the *p*-adic diffusion equation to describe coronavirus’ spread in hierarchically clustered population. This equation has applications to statistical physics and microbiology for modeling *dynamics on energy landscapes*. To move from one social cluster (valley) to another, the virus (its carrier) should cross a social barrier between them. The magnitude of a barrier depends on the number of social hierarchy’s levels composing this barrier. As the most appropriate for the recent situation in Sweden, we consider *linearly increasing barriers*. This structure matches with mild regulations in Sweden. The virus spreads rather easily inside a social cluster (say working collective), but jumps to other clusters are constrained by social barriers. This behavior matches with the covid-19 epidemic, with its cluster spreading structure. Our model differs crucially from the standard mathematical models of spread of disease, such as the SIR-model. We present socio-medical specialties of the covid-19 epidemic supporting our purely diffusional model.

## 1 Introduction

The covid-19 epidemic has many unusual features (see section 2). One of them plays the crucial role in disease spread. We formulate it as the basic assumption of this paper (see also [1]):

**AS0** *Virus’ spread in population is constrained by the hierarchic social cluster structure*.

How can one model mathematically social clustering of population? In a series of works [2]-[7], we constructed *ultrametric clustering* of population by using the system of hierarchically ordered social coordinates and this approach was applied in cognition, psychology, sociology, information theory (see also [8]-[12]). In this paper, we shall use ultrametric diffusion equation [13]-[21] to describe dynamics of coronavirus spread in socially clustered population. It is important to note that ultrametric spaces have treelike geometry, so we shall study dynamics of virus on social trees. To simplify mathematics, consideration is restricted to homogeneous trees with *p*-branches leaving each vertex. Such trees are endowed with algebraic structure, the *p*-adic number fields. We remark that *p*-adic numbers are widely used in number theory and algebraic geometry. Their applications to natural phenomena started with string theory and quantum physics [22]-[24].

The specialties of covid-19 epidemic^1^ are not reflected in the standard mathematical models [25]-[27], such as, e.g., the canonical SIR model [28] and its diffusion-type generalizations, e.g., [29]. Consequently, in spite of the tremendous efforts [30]-[36], mathematical modeling of coronavirus spread cannot be considered as successful. Therefore, we have to search for new mathematical models reflecting better the covid-19 specialties. The recent paper [1] based on **AS0** can be considered as a step in this direction. In it, we studied the problem of *approaching herd immunity in countries like Sweden*, i.e., without lock-down. The coronavirus does not spread throughout population homogeneously as it is described by the standard models of disease spread. Its spread has the clear social cluster character (cf. with disease spread modeling in articles [37]-[40]). The coronavirus spreads relatively easy in a social cluster that was infected by somebody, but approaching other clusters is constrained by social barriers.

Such virus’ spread is described very well by *dynamics on energy landscapes*. The latter is well developed theory with numerous applications to statistical physics (e.g., spin glasses) and microbiology [41]-[47]. An energy landscape is a system of (energy) valleys separated by barriers of different heights having a hierarchic structure. A system (physical, biological) can move inside a valley or jump over a barrier to another valley, with some probability. Thus, the simplest mathematical model is given by *random walks on energy landscapes* (see, e.g., [48]). Behavior of random walking depends crucially of grows of barriers coupled to the hierarchic structure of an energy landscape.

Geometrically the hierarchy of valleys (clusters) of an energy landscape has the treelike structure. As is well known, trees also give the geometric representation for ultrametric spaces and vice verse. Thus, dynamics on energy landscapes, collection of clusters separated by hierarchically ordered barriers, can be represented as dynamics in ultarmetric spaces.

In the first paper [1] on ultrametric approach to disease spread, we explored the random walk in ultrametric spaces [48]. Such random walk is the discrete version of *ultrametric diffusion*. Theory of diffusion equations in ultrametric space is well developed [13]-[21]. In the present paper, we apply its powerful mathematical apparatus for modeling covid-19 spread in hierarchically structured social clusters. The problem of approaching herd immunity is reformulated in terms of ultarmetric diffusion equation. We consider a country without lockdown, but, nevertheless, imposing a bunch of social restrictions (barriers) during the covid-19 epidemic. As the basic example, we consider Sweden (see [1], appendix 2 for compact description of situation in Sweden, March-June 2020, from the viewpoint of imposing social barriers). This reformulation makes the model mathematically rigorous (studies [41]-[47], [48] were at the physical level of rigorousness) and opens the door for development of a variety of new mathematical models of coronavirus spread taking into account the social cluster structure of population.

For Sweden, this problem of approaching herd immunity is of the big value. The country did not impose the lock-down and the system of measures presented by the state epidemiologist Anders Tegnell and his team was aimed to approach herd immunity and, in this way, to make essentially weaker or escape at all the second wave of covid-19 epidemic and may be proceed without vaccination. However, the dynamics of population’s immunity against coronavirus is very slow, essentially slower than it was predicted by Swedish epidemiologists and by mathematical models of disease spread ^2^ (see, e.g., [49]-[51] for reports from Public Health Institute of Sweden, [32]-[34] for attempts of mathematical modeling and [52]-[56] for reports from massmedia).

As we have seen [1], ultrametric random walk (with jumps over mild barriers linearly growing with levels of social hierarchy) generates dynamics with asymptotic behavior of the power type; herd immunity in a social cluster *C* grows as *p*_Im_(*C, t*) = 1 − *t*^−*q*^, *q* > 0. Generally (but, of course, depending on the parameter *q*) this function increases slowly. This asymptotic can explain unexpectedly slow approaching herd immunity by population of Sweden. The basic parameter of the model *q* = *T* log *p/*Δ. Here *T* > 0 is the social analog of temperature, the degree of activity in a society, Δ is the magnitude of the elementary barrier for hopping between nearest social levels. Higher social temperature *T* implies more rapid approaching of herd immunity; higher social barrier Δ implies slower growth of herd immunity.

In the present paper, by using results of work [17] on the relaxation dynamics for diffusion pseudo-differential equation on ultrametric spaces we reproduce the power low for dynamics of herd immunity from [1], for linearly growing barriers. The technique of ultrametric diffusion equations provides the possibility to study this problem for other types of barriers as well as for design of more general mathematical models of covid-19 spread, may be even matching quantitatively with medical data on this epidemic.

## 2 Specialties of covid-19 spread

As was emphasized in introduction, covid-19 epidemic has some specialties. To match these specialties, one has to develop new mathematical models. The fundamental specialty is the social cluster character of coronavirus spread, see **AS0**. Further, we shall discuss a few other virus’ features. They justify the following assumption distinguishing our purely diffusional model of virus spread from the standard SIR-type models:

**AS1** *Intensity of virus’ spreading is relatively insensible to the number of those who have already been infected*.

Now we discuss a few biological and social factors behind this feature of the virus.

- **Covid on surface**. As was shown in study [58], the probability to become infected through some surface (say in a buses, metro, shop) is practically zero. It was found that even in houses with many infected (symptomatic) people, the viruses on surfaces (of say tables, chairs, mobile phones) were too weak to infect anybody. (Their were present, but were not able to infect mouths.)^3^
- **Covid in air**. The virus is neither so much dangerous at the open air, especially if people follow the recommendation to keep 1, 5 m distance between them. In in [58] was pointed out: “The fact that COVID 19 is a droplet infection and cannot be transmitted through the air had previously also been confirmed by virologist Christian Drosten of Berlin’s Charité. He had pointed out in an interview [59] that coronavirus is extremely sensitive to drying out, so the only way of contracting it is if you were to ‘inhale’ the droplets.”
- **Asymptomatic individuals**. As was recently announced [60], WOH collected a lot of statistical data showing that asymptomatic individuals transmit covid-19 virus to other people with very low probability.^4^ At the same time, US Centers for Disease Control and Prevention estimates that about a third of coronavirus infections (35%) are asymptomatic [61]. Hence, about 35% of infected people practically do not contribute in disease spread.
- **No mass-events**. Another important restriction supporting **AS1** is that even in Sweden, mass-events were forbidden, so no public concerts, neither football matches.^5^
- **Superspreaders**. As for many infections, spread of coronavirus has the following feature - the presence of superspreaders of infection. One person can infect really many people. Thus, single person’s contribution in disease spread can be essentially higher than contribution of a few hundreds of usual asymptomatic individuals or many presymptomatic individuals (see more on superspreaders in Appendix).

**AS2** *The number of susceptible people S*(*t*) *is so large comparing with the number I*(*t*) *of those who are infected or the number R*(*t*) *of recovered that we can consider it as constant, S*(*t*) = const, *and exclude it from model’s dynamical equations*.

This assumption implies that for an individual in population under consideration the probability to become infected practically does not depend on the number of recovered. The population is rather far from approaching herd immunity and a disease spreader is surrounded (with the high degree of approximation) by susceptible people. Thus the number of recovered people *R* also can be excluded from dynamics. Of course, this model provdies only the rough picture of the real disease spread, but it reflects the basic features of the covid-19 spread in the states that imposed relatively soft restrictions in relation with epidemic (as, e.g., Sweden, Japan, Belarus).

Denote the probability, for a person from elementary social cluster *x*, to become infected at the instance of time *t* by the symbol *p*_*I*_(*x, t*). To write the evolution equation for *p*_*I*_(*x, t*), we impose the additional assumption:

**AS3** *The distribution of social clusters in the society is uniform: all clusters represented by balls of the same radius have the same measure that is equal to balls’ radius*.

Mathematically **AS3** is formalized through the use of the Haar measure *µ* on **Q**_*p*_. We understand that this is a strong restriction on the social structure of society. But, the main reason for its imposing is just simplification of mathematics. We can consider other distributions on **Q**_*p*_ assigning different weights to social clusters represented by balls of the same radius. (We recall that any point of a ball can serve as its center.)

## 3 Hierarchic treelike geometry of social clusters

We represent the human society as a system of hierarchically coupled (as a treelike structure) disjoint clusters. There are many ways for mathematical modeling of such representations. Theory of *ultrametric spaces* is one of the basic mathematical tools for this purpose. Geometrically ultrametric spaces can be represented as trees with hierarchic levels. Ultrametricity means that this metric satisfies so-called strong triangle inequality:

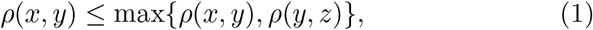

for any triple of points *x, y, z*. Here in each triangle the third side is less or equal not only the sum of two other sides (as usual), but even their maximum. Define balls as usual in metric spaces *B*_*R*_(*a*) = {*x* : *ρ*_*p*_(*x, a*) *≤ R*}, where *a* is a center of the ball and *R* > 0, is its radius. The balls have the following basic properties:

- Two balls are either disjoint or one of them is contained in another.
- Any point of a ball can be selected as its center, i.e., *B*_*R*_(*a*) = *B*_*R*_(*b*) for any *b ∈ B*_*R*_(*a*).

Any ball can be represented as disjoint union of balls of smaller radius, each of the latter can be represented in the same way with even smaller radius and so on. We get hierarchy of balls corresponding to disjoint partitions. Geometrically a ball is a bunch of branches of a tree.

We use the ultrametric balls to represent mathematically social clusters, any cluster is slit into disjoint sub-cluster, each of the latter is split into its own (disjoint) sub-clusters and so on. Inclusion relation generates the hierarchy on the set of social clusters.

In a series of works of the author and his collaborators [2]-[6], ultrametric spaces (geometrically hierarchic trees) were applied for modeling of cognitive, psychological, and social phenomena. This modeling was based on invention of systems of discrete social (or mental in cognitive studies) coordinates *x* = (*x*_*m*_) characterizing (psycho-)social states of individuals. The treelike representation of *social states* is based on selection of hierarchically ordered social factors enumerated by index *m ∈* **Z** = {0, ±1, ±2, …}. (It is convenient to work with coordinates enumerated by integer numbers.) The social importance of coordinates *x*_*m*_ decreases with increase of *m* and increases with decrease of *m*; e.g., social coordinate *x*_0_ is more important than any *x*_*j*_, *j* > 0, but it less important than any *x*_*j*_, *j <* 0. The coordinate *x*_0_ can be considered as a reference point. Depending on context (say socio-economic or socio-epidemic) it can be shifted to the right or to the left. Therefore it is convenient to use positive and negative indexes determining two different directions of social importance of coordinates.

We consider discrete social coordinates, generally, for each *m*, there *N*_*m*_ possible values, *x*_*m*_ = 0, 1, …, *N*_*m*_ − 1, and *N*_*m*_ can vary essentially with *m*. In the treelike representation, numbers *N*_*m*_ determine the number of branches leaving vertexes. Such trees are complicated and we restrict modeling to homogeneous trees for that *N*_*m*_ does not depend on *m*. Moreover, by pure mathematical reasons it is convenient to select *N*_*m*_ = *p*, where *p* > 1 is the fixed prime number. We remark that the corresponding theory was developed even for arbitrary trees (ultrametric spaces), but it is essentially more complicated [18, 19].

Thus, a social state *x* is represented by a vector of the form:

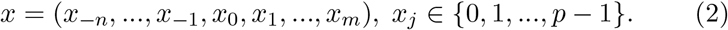

The vector representation of psychical, mental, and social states is very common in psychology and sociology. The essence of our approach [2]- [6] is the hierarchic ordering of coordinates leading to introduction of ultrametric on the state space, see (5).

For our purpose, modeling of epidemic, we can consider, for example, the following hierarchic system of social coordinates; for simplicity, let index *m* = 0, 1, 2, …, so the coordinate *x*_0_ is the most important. It is natural to use it to denote states (e.g., Sweden, Russia, USA,…); *x*_1_ can be used for age; *x*_2_ for chronic diseases, *x*_3_ gender, *x*_4_ for race, *x*_5_ for the town of location, *x*_6_ for the district, *x*_7_ for profession, *x*_8_ for the level of social activity, *x*_9_ for the number of children living with this person, and so on. We understand that such ranking of the basic social factors related to the covid-19 epidemic may be naive and incomplete. The contribution of sociologists, psychologists, and epidemiologists can improve the present model essentially, see even the recent article [57] on mathematical model of evolutionary creation of social types and contribution of genetics and natural selection.

Since the majority of states selected the lock down policy that was not oriented towards approaching herd immunity, we restrict consideration to the Swedish population. So, in the above assigning of social meaning to coordinates they are shifted to the left. We also stress that hierarchy of social factors involved in the covid-19 epidemic can be selected depending on the state, i.e., for each state we create its own system of social clustering coupled to this epidemic. Consider USA, here the population is not so homogeneous with respect to the level of income and the life style connected to income as it is in Sweden. The social factor of belonging to up or low income classes plays the crucial role in covid-19 infecting. It seems that this coordinate should be places as the next (to the right) to the age-coordinate, then the race-coordinate and so on. Thus, the above hierarchy, (age, chronic disease, gender, race, town, district, family,…), that is appropriate for Sweden, should be rearranged for USA, as say (age, income, race, chronic disease, gender, town, district, family,…).^6^

It is convenient to proceed with variable number of coordinates, i.e., not fix *n* and *m*. This gives the possibility to add new coordinates. The space of such vectors can be represented by rational numbers of the form

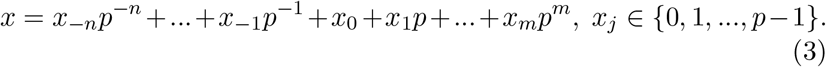

This is the basis of the number-theoretic representation of the space of social states. We shall consider it later. Now we continue in the vector framework.

To use fruitfully ultrametric models, we have to construct a complete metric space. The standard way is approach completeness is to consider infinite sequences of the form:

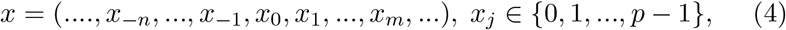

where, for each *x*, there exists *n* such that *x*_−*j*_ = 0, *j* > *n*. Denote the space of such sequences by the symbol **Q**_*p*_. On this space, a metric is introduced in the following way. Consider two sequences *x* = (*x*_*j*_) and *y* = (*y*_*j*_); let *x*_*j*_ = *y*_*j*_, *j < n*, where *n* is some integer, but *x*_*n*_ *≠ y*_*n*_. Then the distance between two vectors is defined as

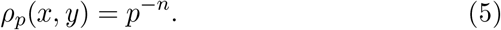

So, if *n* is negative, then distance is larger than 1, if *n* is nonnegative, then distance is less or equal to 1. The *ρ*_*p*_ is an ultrametric. We remark that each ball can be identified with a ball of radius *R* = *p*^*n*^, *n ∈* **Z**. Ball *B*_1_(0) = plays the important role and it is defined by special symbol **Z**_*p*_. As in any ultrametric space, each ball is represented as disjoint union of smaller balls,e.g.,

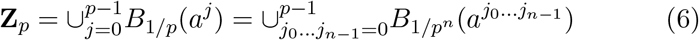

where *a*^*j*^ *∈* **Z**_*p*_ is constrained by condition *x*_0_ = *j* and 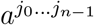 is constrained by conditions *x*_0_ = *j*_0_, …, *x*_*n*−1_ = *j*_*n*−1_, and so on. We recall that in an *ultrametric space, any point of a ball can be selected as its center*.

In our model, *p*-adic balls represent social clusters corresponding to fixing a few social coordinates. For example *C*_*j*_ = *B*_1*/p*_(*a*^*j*^) = {*x ∈***Z**_*p*_ : *x*_0_ = *j*}, in above epidemic coding *C*_*j*_ corresponds to fixing age= *j*; *C*_*ji*_ = *B*_1*/p*_(*a*^*ji*^) = {*x ∈* **Z**_*p*_ : *x*_0_ = *j, x*_1_ = *i*}, age= *j*, gender = *i* for Swedish society or age= *j*, income level= *i* for American society.

Social states, points of **Q**_*p*_, can be cosnidered as balls of zero radius, we call them *elementary social clusters*. Partitions of a ball into disjoint balls of smaller radii corresponds to partition of a social cluster into disjoint subclusters of deeper level of social hierarchy.

Now we turn to the algebraic representation of social states by rational numbers, see (3). The space **Q**_*p*_ endowed with ultrametric *ρ*_*p*_ can be considered as completion of this set of rational numbers and algebraically the elements of **Q**_*p*_ can be represented by power series of the form

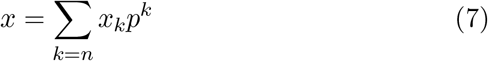

where *x*_*j*_ *∈* {0, 1, …, *p* − 1}, *x*_*n*_ *≠* 0, and *n ∈* **Z**; so only finite number of coordinates with negative index *k* can differ from zero. Such a series converges with respect to ultrametric *ρ*_*p*_. Representation by the power series gives the possibility to endow **Q**_*p*_ with the algebraic operations, addition, subtraction, multiplication, and division (the latter operation is defined only for prime *p*). Hence, **Q**_*p*_ is a *number field, the field of p-adic numbers*. This algebraic representation leads to number-theoretic representation of ultrametric, *ρ*_*p*_(*x, y* = |*x* − *y*|_*p*_, where *x →* |*x*|_*p*_ is the *p*-adic analog of the real absolute value; per definition, for *x* given by series (7),

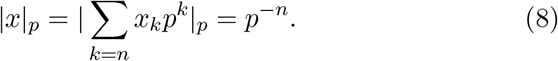

It satisfies the strong triangle inequality playing the fundamental role in *p*-adic analysis:

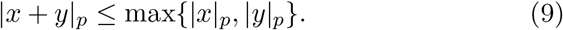

## 4 Modeling the virus spread with ultrametric diffusion equation

An elementary social cluster given by a point of **Q**_*p*_ is a mathematical abstraction. Real clusters are represented by balls of finite radii. Therefore it is interesting to study the evolution of average probability for cluster 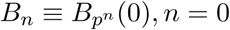, ±1, ±2, Under assumption **AS3**, this quantity is represented as the integral with respect to the Haar measure:

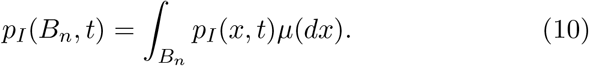

Under the above assumptions on the social structure of population and its interaction with the virus (including restrictions imposed by authorities in connection with epidemic), we can write the following master equation for probability *p*_*I*_(*x, t*),

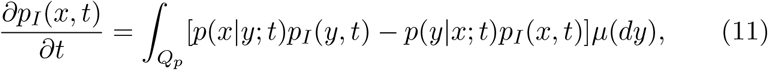

where *p*(*x*|*y*; *t*) is the transition probability: the probability that the virus being present in (elementary) cluster *y* would jump to cluster *x*. We suppose that this probability does not depend on time *t* and it is symmetric, i.e., *p*(*x*|*y*) = *p*(*y*|*x*). Under these assumptions, the master equation has the form

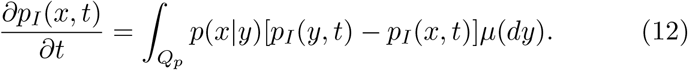

It is natural to assume that the transition probability decreases with increasing of the distance between two clusters, for example, that

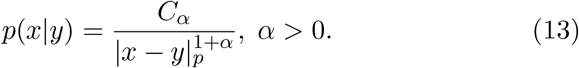

Here *C*_*α*_ > 0 is a normalization constant, by mathematical reasons it is useful to select distance’s power larger than one. Hence,

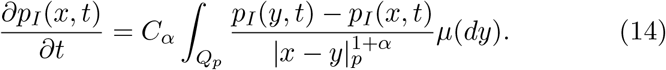

The integral operator in the right-hand side is the operator of fractional derivative *D*^*α*^ (the Vladimirov operator), see [13]. Thus, the dynamics of the probability to become infected for those belonging to an elementary social cluster is described by *the p-adic diffusion equation:*

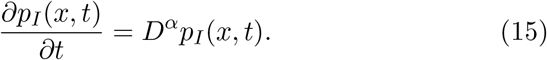

To formulate the Cauchy problem, we have to add some initial probability distribution. We select the uniform probability distribution concentrated on a single ball, initially infected social cluster *B*_*n*_,

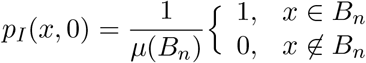

This equation and its various generalizations were studied by many authors, for applications to physics and biology and by pure mathematical reasons, see, e.g., [13]. We are interested in the relaxation regime, i.e., asymptotic of average probability *p*_*I*_(*B*_*n*_, *t*) for large *t*. We use the mathematical result from [17] (see also [18, 19]) and obtain that the average probability has the power behavior:

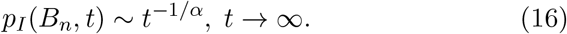

Thus the average probability to become infected in a social cluster decreases rather slowly with time. If *α* >> 1, i.e., the virus transition probability decreases very quickly with increase of the distance between social clusters, then *p*_*I*_(*B*_*n*_, *t*) decreases very slowly with time, it is practically constant. If 0 *< α <<* 1, so the virus transition probability decreases relatively slowly with increase of the distance, then *p*_*I*_(*B*_*n*_, *t*) decreases sufficiently quickly with time. We shall discuss these behaviors in section 5 by assigning bio-social meaning to the parameter *α*.

We present some graphs corresponding to different values of *α* at Fig. 1.

**Figure 1:**
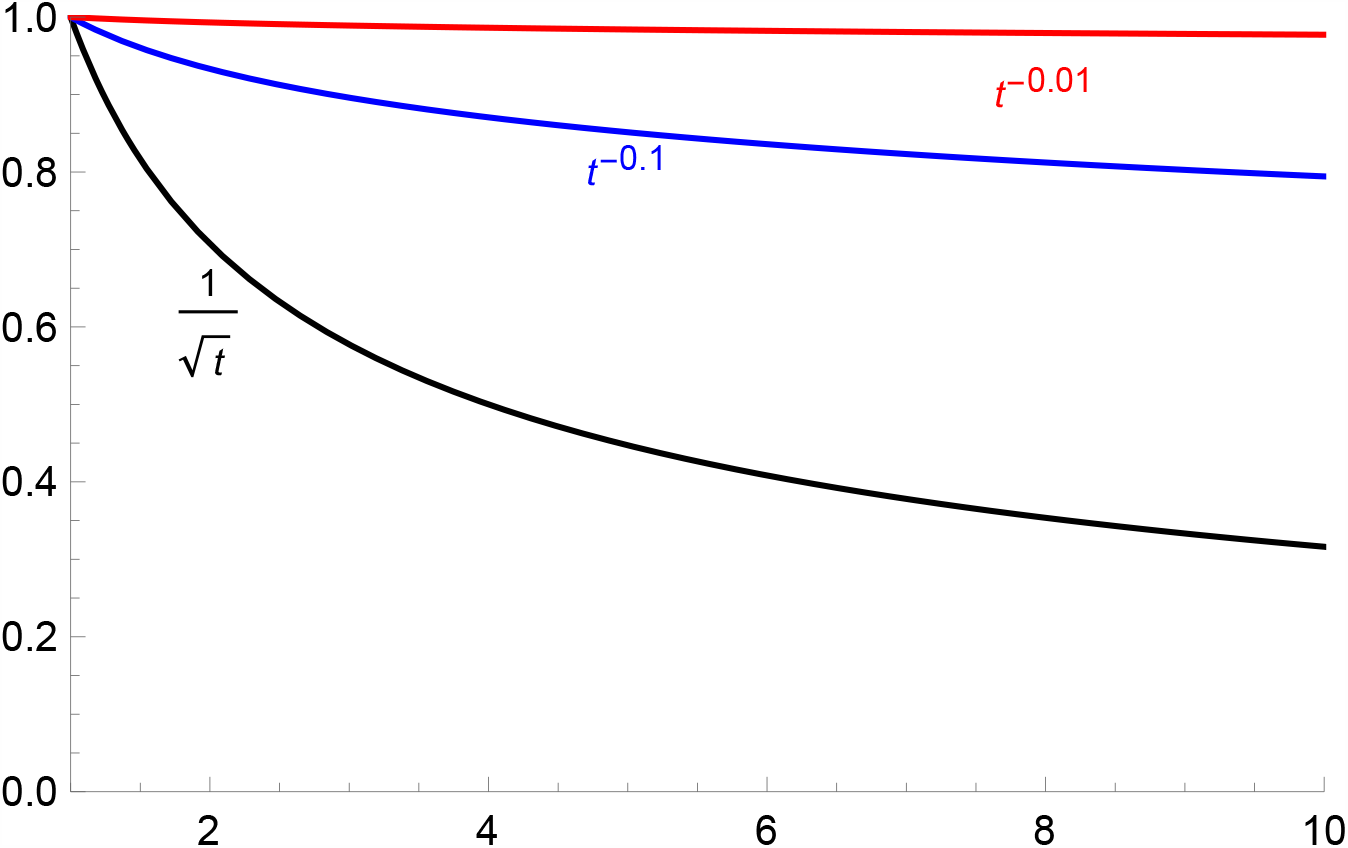
Asymptotic behavior of probability to become infected. (For fixed social temperature *T*, the upper graphs correspond to one-step barrier growth 10 and 100 times, respectively.

Consider now a kind of “integral immunity”, combination of innate and adaptive components, defined as the probability of not become infected:

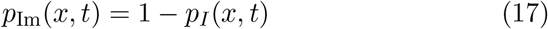

and its average over social cluster represented by ball *B*_*n*_,

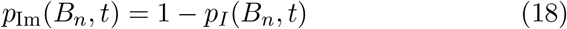

This function increases relatively slowly with time, see Fig. 1. Its asymptotic behavior depends on the parameter *α* determining how rapidly the transition probability between social clusters decreases with increase of the distance between them.

## 5 Virus’ random walk on the hierarchic social tree

The mathematical result on the relaxation regime for the *p*-adic diffusion [17] is generalization of studies on random walks on ultrametric spaces describing dynamics on energy landscapes [48]-[47]. There are given energy barriers Δ_*m*_ separating valleys, movement from one valley to another valley is constrained by necessity to jump over a barrier between them. This random walk model gives a good heuristic picture of the virus spread, as jumping from one social cluster (valley) to another, where clusters (valleys) are separated by social barriers (mountains) of different heights. Geometrically such random walk is represented as jumps on a tree between the levels of social hierarchy. Our model (selection of the transition probability in the form (13)) corresponds to *barriers growing linearly with the number of elementary jumps*. The relaxation regime of the power form is obtained for the number of hierarchy’s levels approaching infinity, i.e., for ideal trees with infinitely long branches, as ultrametric spaces they are represented by **Q**_*p*_.

The virus plays the role of a system moving through barriers in models of dynamics on energy landscapes (see [48], [41]-[47] and references herein). In our case, these are social barriers between social clusters of population. The virus performs a complex random walk motion inside each social cluster moving in its sub-clusters, goes out of it and spreads through the whole population, sometimes the virus comes back to the original cluster from other social clusters that have been infected from this initial source of infection, and so on. During this motion the virus should cross numerous social barriers.

Instead of virus walking through the social tree, we can consider a person. A person of the social type *x* can interact with persons of other social types. The temporal sequence of social contacts of some persons can have a very complicated trajectory, visiting numerous clusters (but the probability of approaching a cluster depends crucially on social barriers).

Let virus encounters a barrier of size Δ_*m*_, in hopping a distance *m* (crossing *m* levels of hierarchy), where Δ_1_ *<* Δ_2_ *<* … *<* Δ_*m*_ *<* It is supposed that barriers Δ_*m*_ are the same for all social clusters, i.e., they depend only on distance, but not on clusters. This assumption reflects a kind of epidemic égalit’e of all social groups, the barriers preventing spread of the virus that are imposed by state authorities are the same for all social groups.

Consider the tree at Fig. 3. We identify the lengths of branches between vertexes with magnitudes of barriers. Then the barriers on this tree depend on clusters, so from this viewpoint the social tree is not homogeneous.

**Figure 2:**
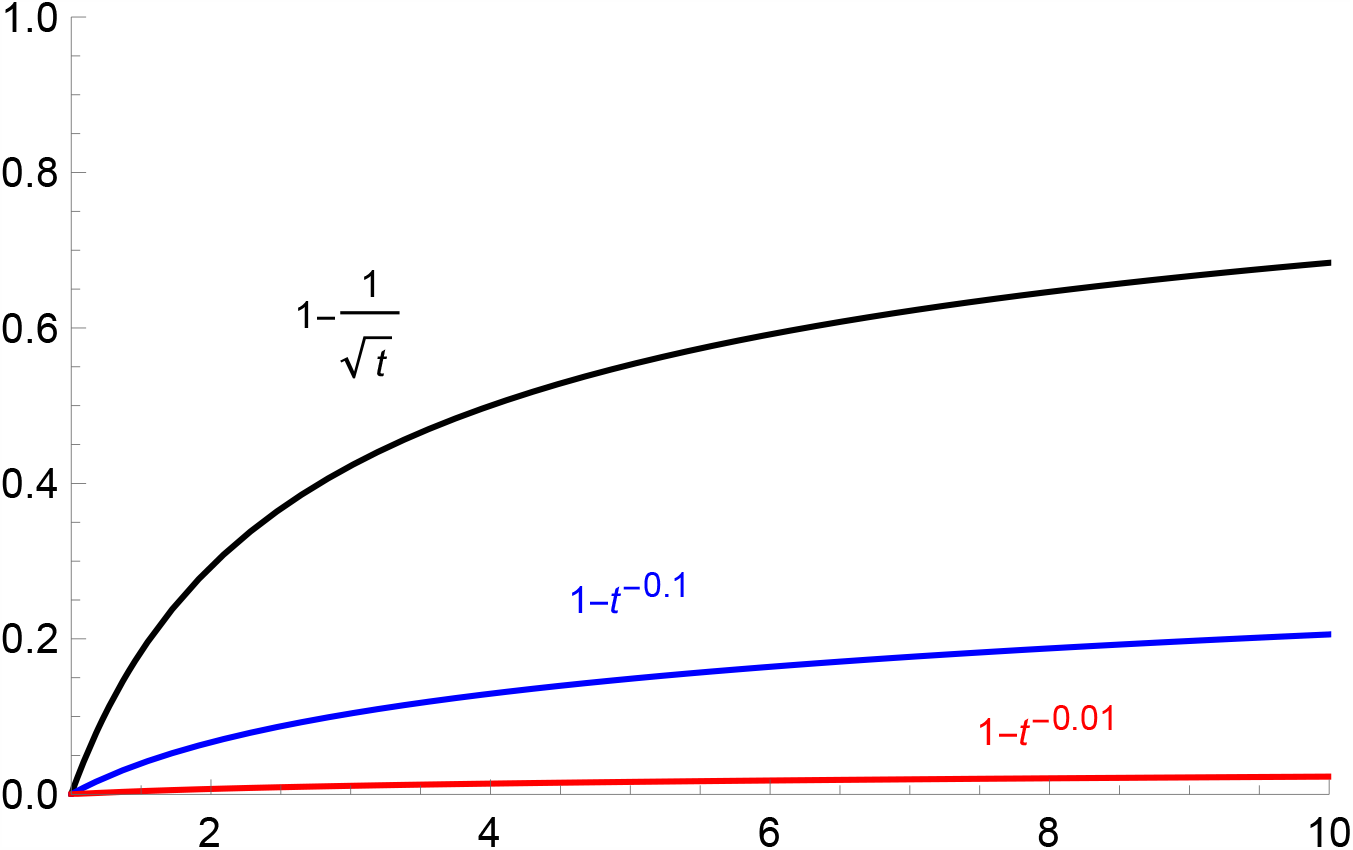
Asymptotic behavior of probability to become immune; increasing of herd immunity (for fixed social temperature *T*, the upper graphs correspond to one-step barrier growth 10 and 100 times, respectively.

**Figure 3:**
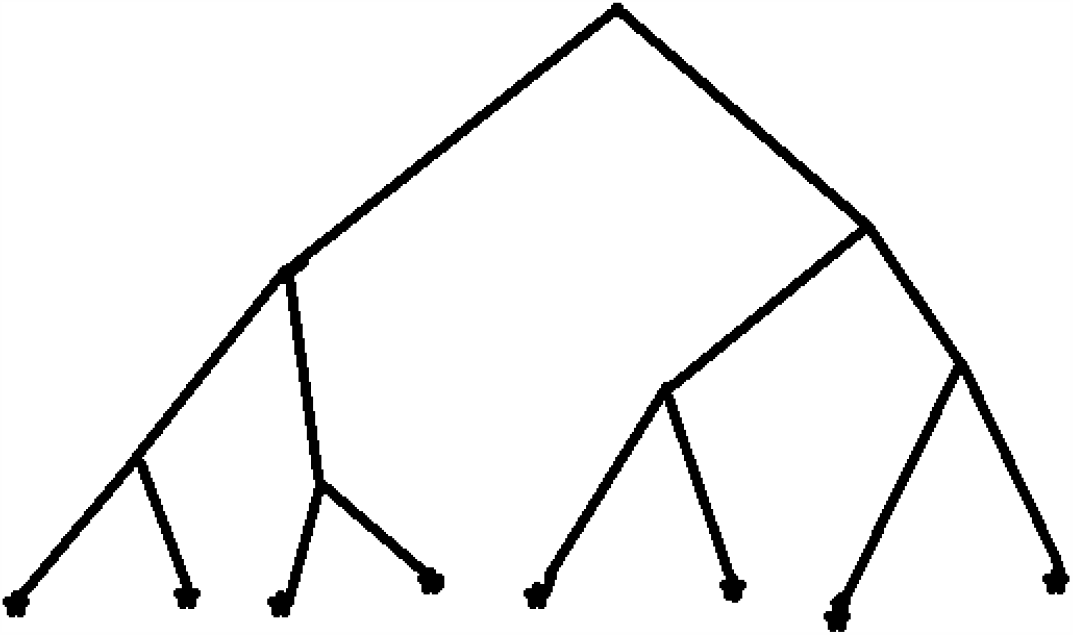
Treelike configuration space

Consider the energy landscape with a uniform barrier Δ, at every branch point; that is, a jump of distance 1 involves surmounting a barrier Δ, of distance 2, a barrier 2Δ, and so on. Hence, barriers linearly grow with distance *m*,

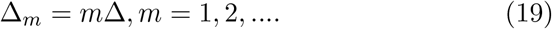

It seems that this type of behavior is the most natural from the viewpoint of social connections during the covid-19 epidemic in Sweden. Barriers are sufficiently high, but they still are not walls as during the rigid quarantine (as say in Italy, France, or Russia). For such linearly increasing barriers one can derive the following asymptotic behavior (17) of the relaxation probability [48], where in physics and biology the parameter

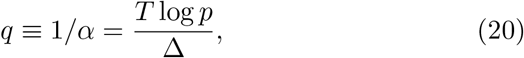

Here the new parameter *T* has the meaning of temperature. Thus behavior of distance between valleys of the energy landscape is determined by the size of the barrier for one-step jump Δ and temperature. We rewrite formula (13) for transition probability by using these parameters:

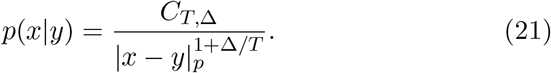

In our model, we introduce the notion of *social temperature T*. As in physics, this parameter calibrates energy, in our case social energy. The latter represents the degree of social activity, the magnitude of social actions. Although the notions of social temperature and energy are not so well established as in physics, they can be useful in socio-physical modeling (see [63] and references herein, starting with the works of Freud and Jung). Probability that the virus jumps from the elementary social cluster *y* to another cluster *x* grows with growth of social temperature. For high *T*, virus (or its spreader) easily move between social clusters. If *T <<* 1, the infection is practically confined in clusters. If barrier Δ increases for the fixed parameter *T*, then the transition probability decreases and vice verse.

Starting with expression (21), we obtain the relaxation asymptotic in the form:

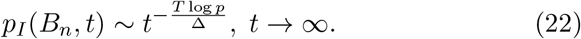

Thus, for large *t*, the average probability to become infected in social cluster *B*_*n*_ decreases quicker with increase of social temperature *T*. Decrease of the one-step jump barrier Δ implies the same behavior.

Immunity probability *p*_*Im*_(*B*_*n*_, *t*) behaves in the opposite way. It increases quicker with increase of social temperature and decrease of the social barrier Δ.

## 6 Concluding remarks

In this paper, we continue development of a new mathematical model of disease spread reflecting specialties of covid-19 epidemic. We especially emphasize the social cluster character of the virus spread, **AS0**. Such clustered spread can be modeled with dynamical systems on ultrametric spaces. Social clusters are represented by ultrametric balls. The basic feature of ultrametric balls is that they are either disjoint or one is included in another. This is the root of a the hierarchic structure of an ultrametric space. Geometrically ultrametric spaces are represented by trees with balls given by bunches of branches with the common root.

In this paper, we model the dynamics of coronavirus with ultrametric diffusion equation^7^, its simplest version corresponding to *p*-adic trees and linearly increasing social barriers. Asymptotic of probability *p*_Im_(*t*) to become immune against the virus is presented at Fig. 2. Generally, it increases slowly, the speed of increasing depends on the basic parameter of the model *q* = *T* log *p/*Δ.

In a society with low social temperature and high barriers between social clusters, *p*_Im_(*t*) increase so slowly that there is practically no hope to approach herd immunity.

## Data Availability

no data

## Appendix

Superspreader is an unusually contagious individual who has been infected with disease; someone who infected the number of people far exceeding the two to three. As was pointed out in MIT Technology Review [62]: “For covid-19, this means 80% of new transmissions are caused by fewer than 20% of the carriers – the vast majority of people infect very few others or none at all, and it is a select minority of individuals who are aggressively spreading the virus. A recent preprint looking at transmission in Hong Kong supports those figures, while another looking at transmission in Shenzhen, China, pegs the numbers closer to 80/10. Lots of outbreaks around the world have been linked to single events where a superspreader likely infected dozens of people. For example, a choir practice in Washington State infected about 52 people; a megachurch in Seoul was linked to the majority of initial infections in South Korea; and a wedding in Jordan with about 350 guests led to 76 confirmed infections.” The bad news is that, for the moment, we cannot identify diagnostically superspreaders.

## Footnotes

1 See section 2, “covid on surface”, “covid in air”, “asymptomatic individuals”, “no mass-events”, “superspreaders”; in this paper we are interested in mild restrictions, as in Sweden, i.e., without lock-down.

2 In particular, by models Tom Britton [32, 33] that was used by Swedish State Health Authority predicted that herd immunity will be approached already in May; Anders Teg-nell also announced, starting from the end of April 2020, that Sweden would soon approach herd immunity, but it did not happen, neither in May, nor June and July.

3 Mr Streeck, a professor for virology and the director of the Institute of virology and HIV Research at the University Bonn, clarified [58]: “There is no significant risk of catching the disease when you go shopping. Severe outbreaks of the infection were always a result of people being closer together over a longer period of time, for example the apré-ski parties in Ischgl, Austria.” During extended and careful study in Heidelberg (the German epicenter of the covid-19 epidemic) his team could also not find any evidence of living viruses on surfaces. “When we took samples from door handles, phones or toilets it has not been possible to cultivate the virus in the laboratory on the basis of these swabs. … To actually ‘get’ the virus it would be necessary that someone coughs into their hand, immediately touches a door knob and then straight after that another person grasps the handle and goes on to touches their face.” Streeck therefore believes that there is little chance of transmission through contact with so-called contaminated surfaces.

4 ”We have a number of reports from countries who are doing very detailed contact tracing. They’re following asymptomatic cases, they’re following contacts and they’re not finding secondary transmission onward. It is very rare – and much of that is not published in the literature,” Van Kerkhove, WOH official said on June 6, 2020. “We are constantly looking at this data and we’re trying to get more information from countries to truly answer this question. It still appears to be rare that an asymptomatic individual actually transmits onward.” [60].

5 In Sweden, restaurants and night clubs were open, but such events were not of mass-character. The presence in a night club or in a restaurant of one infection spreader has practically the same impact as say 5 spreaders, the effect of closed space. Moreover, the distance between the tables in restaurants also diminished the effect of high number of infected in the population. During the intensive phase of the covid-19 epidemic (the end of March and April 2020) restaurants terminated self-serving during lunches (so typical in Sweden).

6 Income did not play any role in Sweden during the covid-19 epidemic.

7 The use of purely diffusional model is supported by specialties of covid-19 epidemic, presented in section 2. Of course, this model is only approximate. But, it seems that it gives the right asymptotic of probabilities, to become infected and immune, in socially clustered society.

